# Informing Variant Assessment using Structured Evidence from Prior Classifications (PS1, PM5, and PVS1 Sequence Variant Interpretation Criteria)

**DOI:** 10.1101/2022.05.16.22275073

**Authors:** Vineel Bhat, Ivan A. Adzhubei, James D. Fife, Matthew Lebo, Christopher A. Cassa

## Abstract

**Purpose:** To explore whether evidence of pathogenicity from prior variant classifications in ClinVar could be used to inform variant interpretation using the ACMG/AMP clinical guidelines.

**Methods:** We identify distinct SNVs which are either similar in location or in functional consequence to pathogenic variants in ClinVar, and analyze evidence in support of pathogenicity using three interpretation criteria.

**Results:** Thousands of variants, including many in clinically actionable disease genes (ACMG SFv3.0), have evidence of pathogenicity from existing variant classifications, accounting for 2.5% of non-synonymous SNVs within ClinVar. Notably, there are many variants with uncertain or conflicting classifications which cause the same amino acid substitution as other pathogenic variants (PS1, N=323), variants which are predicted to cause different amino acid substitutions in the same codon as pathogenic variants (PM5, N=7,692), and LOF variants which are present in genes where many LOF variants are classified as pathogenic (PVS1, N=3,635). The majority of these variants have similar computational predictions of pathogenicity and splicing impact as their associated pathogenic variants.

**Conclusion:** Broadly, over 1.4 million SNVs exome-wide could make use of information from previously classified pathogenic variants. We have developed a pipeline to identify variants meeting these criteria, which may inform interpretation efforts.

## Introduction

Assessing the clinical validity of variant classifications is a major goal in precision medicine.^1^ While diagnostic sequencing has advanced dramatically, the interpretation of monogenic variants remains challenging, even in established disease genes, often on the basis of their low frequencies, lack of observations in clinical cases, and/or availability of functional data.^2,3^ Because of the inherent complexities in variant interpretation and the value of many sources of evidence, the American College of Medical Genetics and Genomics (ACMG) and the Association for Molecular Pathology (AMP) published clinical guidelines for sequence variant interpretation (SVI) in 2015.^2^ The SVI guidelines have provided opportunities to standardize and increase the consistency of variant assessment across clinical laboratories, increasing the utility of reported findings in publicly accessible databases.^6,7^ These guidelines describe sources of evidence and criteria which would help classify a variant as pathogenic or benign, and have been further defined for many criteria.^8–14^ When there are no or few reports describing an identified variant, labs must rely primarily on these sources of evidence, including computational predictive techniques, functional assays, and published gene-level evidence to predict functional effect.^6^

The clinical knowledge base of disease variants is growing rapidly: there are over 460,000 non-synonymous single nucleotide variants (nsSNVs) which have been reviewed by diagnostic labs and deposited to ClinVar.^15^ Among these variants, a majority are classified as VUS (66.4%), have conflicting interpretations (5.7%), or have no assertion provided (1.3%), accounting for 73.4% of variants which do not have clear interpretations.^16^ A recent study found that fewer than 1% of VUS were reclassified in ClinVar over a 3.5 year period, which demonstrates that this problem is being resolved very slowly.^17^ These conflicting or uncertain assessments limit opportunities to improve clinical management and, in rare instances, have been associated with overtreatment (*e*.*g*., unnecessary surveillance or prophylactic surgery).^18,19^

Here, we evaluate the extent to which information from prior variant classifications might be used to inform assessments of existing or novel variants. We make use of variant classifications which have been previously submitted to ClinVar with pathogenic or likely pathogenic (P/LP) assertions, which can provide evidence of pathogenicity for other variants which are similar in location or functional consequence. Specifically, we evaluate how frequently multiple variants affect the same codon (whether causing the same amino acid substitution or not), and also evaluate how frequently loss-of-function (LOF) variants are present in genes where LOF variants are known to be pathogenic. We note that all submissions are being correctly processed by ClinVar and diagnostic labs may have intentionally classified a variant as VUS even if there is sufficient evidence to classify it as P/LP from the clinical guidelines.

## Materials and Methods

### Variant inclusion criteria and annotation

We analyze all nsSNVs in ClinVar^15^ and in dbNSFP^20^, which are filtered and annotated as described in the supplementary methods. When analyzing clinical assessments from ClinVar, our P/LP category includes ‘Pathogenic’, ‘Likely pathogenic’, and ‘Pathogenic/Likely pathogenic’ classifications, and B/LB includes ‘Benign’, ‘Likely benign’, and ‘Benign/Likely benign’ classifications. Importantly, we use the term VUS inclusively to capture all variants with an inconclusive classification, including ‘Uncertain significance’, ‘Conflicting interpretations of pathogenicity’, or ‘not provided’.

### Evaluation of ACMG/AMP interpretation criteria

We evaluate predictive evidence for three ACMG/AMP interpretation criteria:

- **Pathogenic Strong 1 - PS1:** The interpretation guidelines specify that a variant which leads to the same amino acid substitution as a pathogenic variant has strong evidence of pathogenicity, as it would be expected to lead to the same change in protein structure or function.
- **Pathogenic Moderate 5 - PM5:** The interpretation guidelines specify that a variant which leads to a different amino acid substitution in the same codon as a pathogenic variant has moderate evidence of pathogenicity, as changes to the same protein codon may be similarly impactful in protein structure or function.
- **Pathogenic Very Strong 1 - PVS1:** The interpretation guidelines specify that a variant which is expected to lead to loss-of-function in a gene where loss-of-function variants are known to be pathogenic has very strong evidence of pathogenicity.

For these three criteria, we analyze how often a variant which is annotated as uncertain, conflicting, or not provided can benefit from knowledge of variants with existing clinical classifications (**Figure 1a**). For the PS1 interpretation criterion, we identify how often two variants which lead to the same amino acid substitution, including nonsense variants, carry inconsistent classifications within ClinVar, and also analyze the frequency at which two VUS encode the same change. For the PM5 criterion, we identify how often two variants which lead to distinct amino acid substitutions in the same codon, excluding nonsense variants, carry inconsistent classifications within ClinVar. Finally, for PVS1 evidence, we make use of LOFTEE to identify variants which might not be expected to impact function (*e*.*g*., present in the last 50 bp, low expected splicing impact, or within NAGNAG sites)^21^. LOFTEE labels variants which pass all filtering criteria as ‘High Confidence’. We restrict our analysis to LOF which are described in ClinVar as VUS and are present in genes where greater than 50% of LOFTEE ‘High Confidence’ variants are classified as pathogenic.

**Figure 1:**
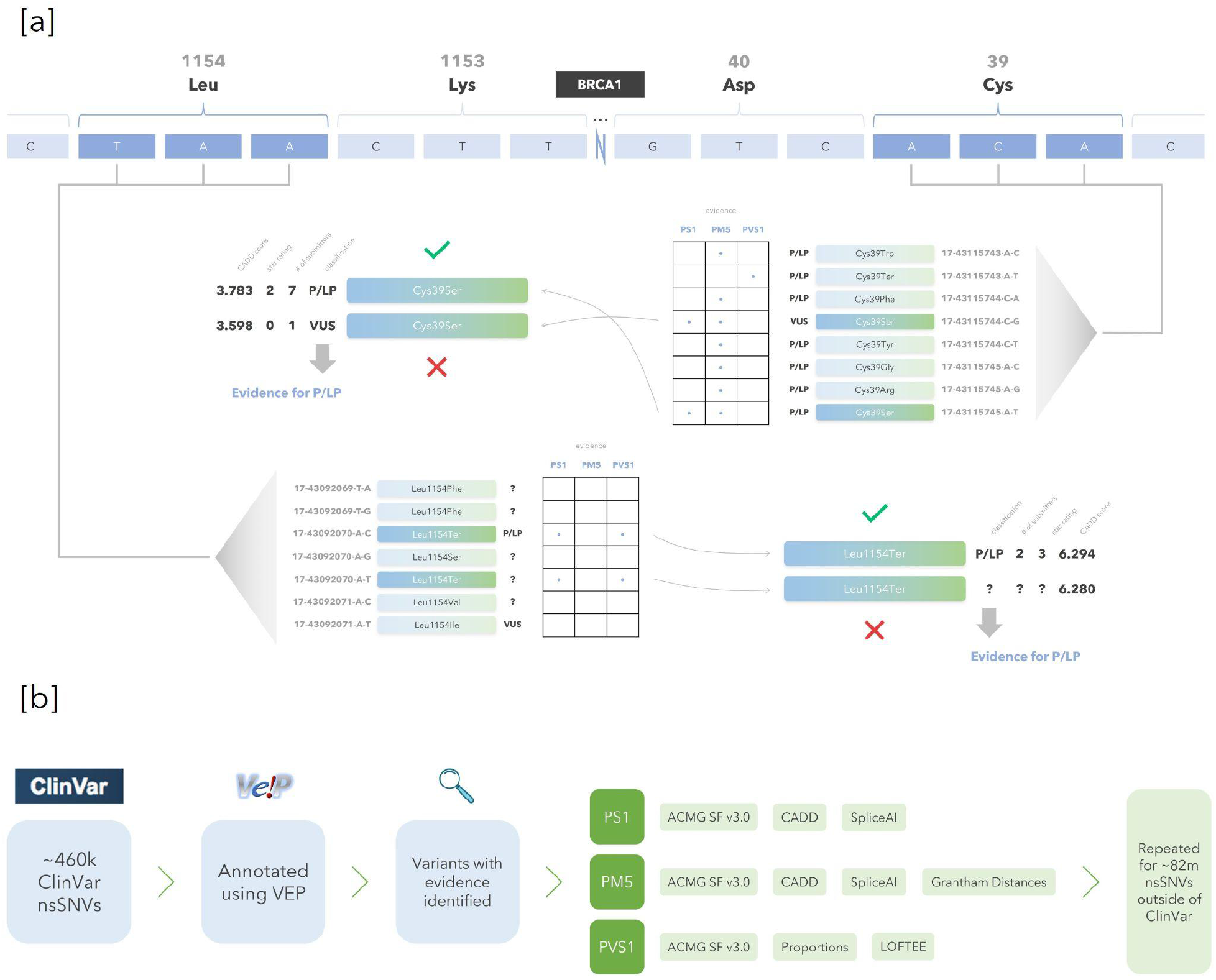
Examples of evidence in favor of pathogenicity in *BRCA1* and methods flow diagram. **[a]** Examples of evidence in favor of pathogenicity in *BRCA1*. ‘PS1’ evidence applies to a variant which encodes the same amino acid substitution as a variant previously classified as pathogenic, ‘PM5’ evidence applies to a variant which encodes a distinct amino acid substitution in same codon as a variant previously classified as pathogenic, and ‘PVS1’ evidence applies to a LOF variant which is in a gene where LOF variants are known to be pathogenic. In the first example, variant ‘17-43115745-C-G’ within codon 35 in *BRCA1* is classified as VUS, has strong evidence of pathogenicity (PS1) based on the P/LP classification of variant ‘17-43115744-A-T’ which encodes the same amino acid substitution, as well as moderate evidence of pathogenicity (PM5) from several other variants within the same codon. In the second example, variant ‘17-43092070-A-T’ within codon 1154 in *BRCA1* does not have a classification in ClinVar, and has strong evidence of pathogenicity (PS1) based on the P/LP classification of variant ‘17-43092070-A-C’ which encodes the same amino acid substitution. The variant also has very strong evidence of pathogenicity (PVS1) as it is a LOF variant and LOF variants are frequently assessed as pathogenic in *BRCA1*. In both examples, the VUS or variant outside of ClinVar with evidence of pathogenicity has a CADD score similar to its P/LP partner which encodes the same amino acid substitution. **[b]** Methods flow diagram. ClinVar variants are annotated and filtered to identify variants which have PS1, PM5, or PVS1 evidence. Multiple tools are used to analyze each evidence tier, and the same process is repeated for the ∼82 million variants exome-wide which are outside of the ClinVar database.

Additionally, we evaluate these criteria for all non-synonymous SNVs outside of ClinVar. We also consider whether previously classified benign variants may provide evidence in support of benign classification, though the ACMG/AMP does not describe analogous criteria for this case.

### Measuring the similarity of variant pairs

We use the term ‘variant pair’ to describe two distinct SNVs which are related to one another because they alter the same protein codon. We measure the extent to which variant pairs have similar computationally predicted impacts, experimental functional assay values, and allele frequencies. For each variant pair, we compare CADD^22^ scores of predicted functional impact, SpliceAI^23^ scores of splicing impact, and functional data in *BRCA1*^24^ for PS1 and PM5 criteria. For the PVS1 criterion, we analyze computational predictions for loss-of-function SNVs (nonsense, canonical splice acceptor, and canonical splice donor variants) using LOFTEE classifications and flags^21^. For all three criteria, we evaluate whether VUS are in clinically actionable disease genes (ACMG SF v3.0) and provide supporting evidence which may be used to prioritize variants for reassessment (**Figure 1b**).

## Results

### Distinct sequence variants which lead to the same amino acid substitution

Within ClinVar, we find many instances where the same amino acid substitution is encoded by more than one distinct sequence variant – in total 7,639 SNVs encoding 3,770 unique amino acid substitutions. Among the codons, 1,728 have at least one variant which has been previously classified as P/LP of which 81.3% (N=1,405) are classified exclusively as P/LP, while 18.4% (N=318) contain variants classified as both P/LP and VUS, and 0.3% (N=5) contain variants classified as both P/LP and B/LB (**Table 1**). There are no codons in which P/LP, B/LB, and VUS variants are present.

**Table 1:**
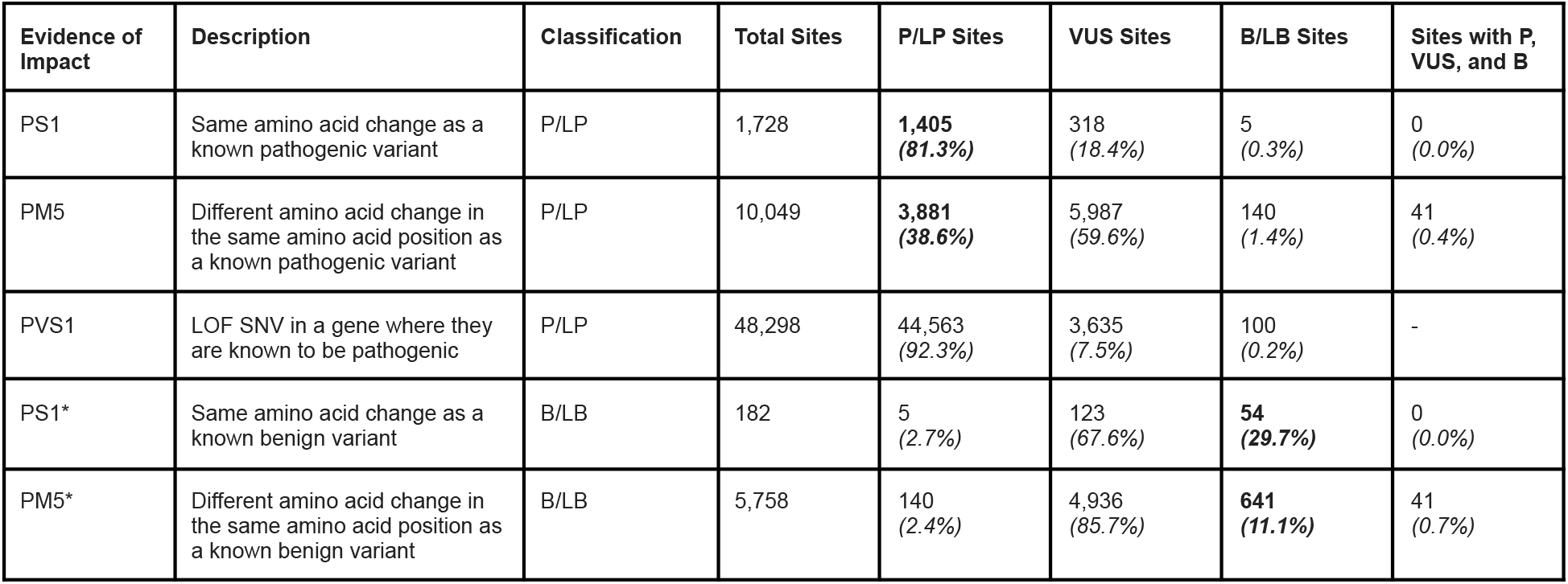
Number of unique sites in ClinVar by evidence and classification. For PS1 and PM5, sites refer to unique codons, while for PVS1, they refer to unique variants. In the case of PS1 and PM5 specifically, P/LP, VUS, and B/LB sites represent codons where only variants with that classification exist in the presence of pathogenic or benign variants within the codon (*e*.*g*., P/LP PS1 Unique B/LB Sites are codons in which at least one P/LP variant and one B/LB variant encode the same amino acid substitution, but no VUS exist in the same codon). Bolded values highlight the number of codons which only have variants of a certain classification. ^*a*^Form of evidence not described in ACMG/AMP guidelines.

Overall, this translates to hundreds of unique ClinVar VUS which have strong evidence of pathogenicity (PS1), and information on their clinical significance and review status values are provided (**Supplementary Table 1, Supplementary Table 2**). Among the VUS, we find 212 unique ClinVar submissions across 116 variants within the genes specified for review of secondary findings (ACMG SF v3.0, 72 genes excluding *HFE*, as p.Cys282Tyr was not part of any set)^25^. This could result in hundreds of patients receiving clinically actionable information related to updated variant classifications.

We further analyze these ‘P/LP-VUS variant pairs’ (P/LP and VUS variants which encode the same amino acid substitution, N=333, 80.2% missense / 19.8% nonsense) to determine whether each VUS is expected to have a similar functional effect as its related P/LP variant. We find that the VUS largely appear to have similar computational predictions of functional effects when compared to their pathogenic partners. When comparing CADD scores for the PS1 criterion, we find very strong correlation (R^2^=0.942, MSE=0.223, y=0.997x, **Figure 2a**) with a mean difference of 0.24 (σ = 0.41). Additionally, we find no significant difference between the CADD scores of P/LP and VUS variants (Mann-Whitney U P=0.980).

**Figure 2:**
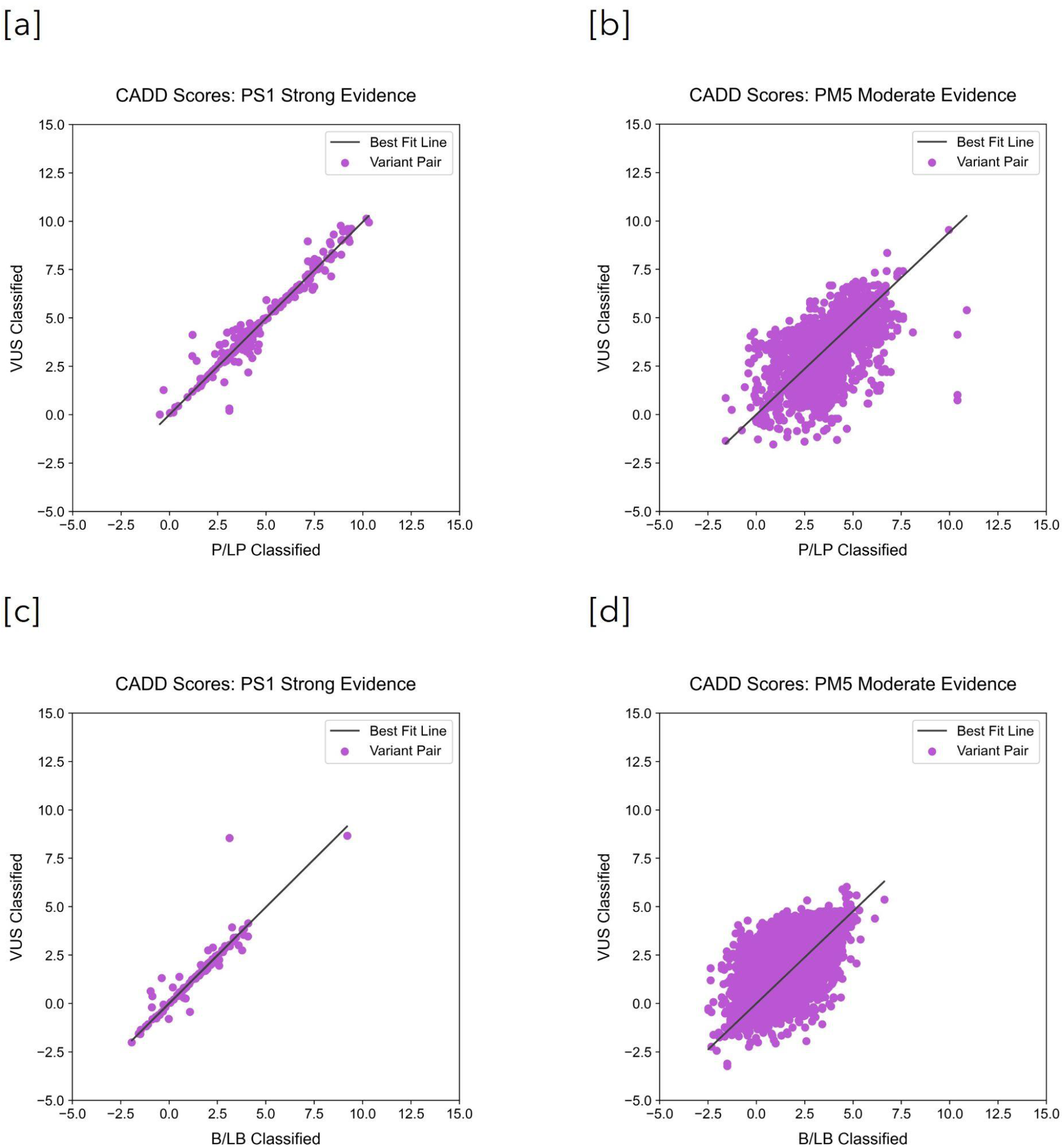
CADD scores of variant pairs are correlated. Individual points represent variant pairs where one variant has a known annotation (P/LP or B/LB) and there is a related VUS affecting the same codon. Each dot compares the CADD score of the variant with an annotation to that of the related VUS. **[a]** CADD scores for P/LP-VUS PS1 variant pairs. The variant pairs have highly correlated scores (R^2^=0.942) and are not significantly different (Mann-Whitney U p=0.980). **[b]** CADD scores for P/LP-VUS PM5 missense variant pairs. The variant pairs are correlated (R^2^=0.169). **[c]** CADD scores for B/LB-VUS PS1 variant pairs. The variant pairs have highly correlated scores (R^2^=0.860) and are not significantly different (Mann-Whitney U p=0.994). **[d]** CADD scores for B/LB-VUS PM5 missense variant pairs. The variant pairs are correlated (R^2^=0.376).

Two distinct sequence variants could cause the same amino acid substitution yet have different splicing effects (*e*.*g*., the P/LP variant has a splicing effect while the VUS does not). To determine whether this may be a primary driver of discordant variant classifications observed in the PS1 variant pairs, we analyze the splicing impact for each variant using SpliceAI^23^. We find that this is unlikely to be the explanation in 98.8% of discordant P/LP-VUS variant pairs (**Figure 3a**). Among 329 distinct P/LP-VUS pairs with SpliceAI annotations, only 0.6% (N=2) of the P/LP variants were predicted to have a high impact on splicing (score >0.8) when their VUS partners were predicted to have a weak or low impact on splicing (score <= 0.5), and only 0.6% (N=2) of the P/LP variants were predicted to have an intermediate impact on splicing (score (0.5, 0.8]) when their VUS partners were predicted to have a weak or low impact.

**Figure 3:**
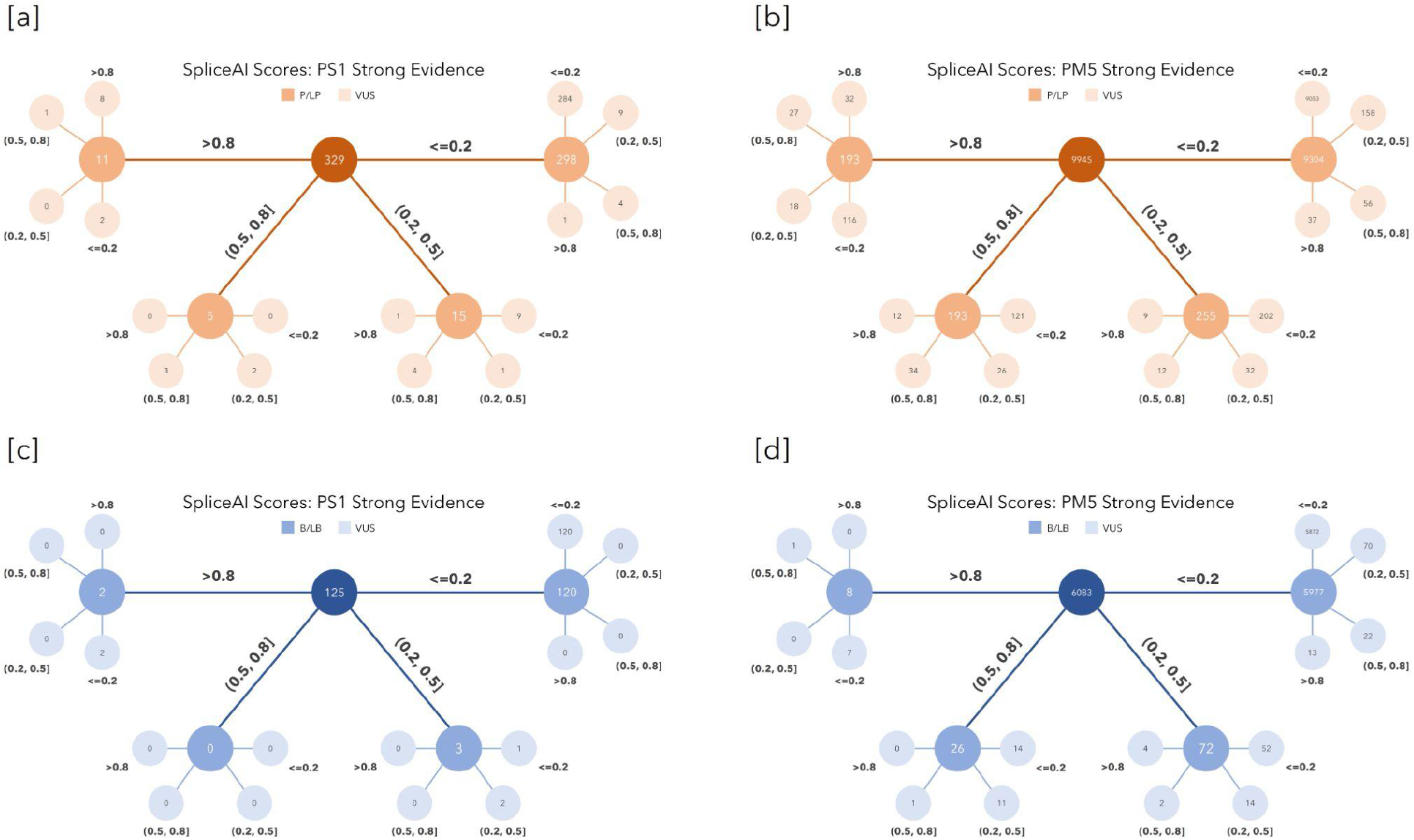
SpliceAI scores of related variants. To evaluate how frequently variants with discordant classifications may be attributable to different splicing impacts, we use SpliceAI. Score ranges of >0.8, >0.5 and <=0.8, >0.2 and <=0.5, and <=0.2 represent high, intermediate, weak, and low predicted impacts on splicing, respectively. **[a]** SpliceAI scores for P/LP-VUS PS1 variant pairs. In 86.3% of pairs (N=284), both variants have low predicted splicing impact. **[b]** SpliceAI scores for P/LP-VUS PM5 variant pairs. In 91.0% of pairs (N=9,053), both variants have scores <=0.2. **[c]** SpliceAI scores for B/LB-VUS PS1 variant pairs. In 96.0% of pairs (N=120), both variants have scores <=0.2. **[d]** SpliceAI scores for B/LB-VUS PM5 variant pairs. In 96.5% of pairs (N=5,872), both variants have scores <=0.2.

Applicability to benign classifications: While there is no ACMG/AMP guideline which is analogous to the PS1 criterion for benign variation, we measured the extent to which the same principles could potentially provide interpretation evidence. Similar to evidence in favor of a pathogenic classification, VUS-B/LB variant pairs (two distinct sequence variants leading to the same amino acid substitution with one B/LB and one VUS classification [in the absence of P/LP variants in the same codon], N=126, 99.2% missense / 0.8% nonsense) show strong correlation in CADD scores (R^2^=0.860, MSE=0.366, y=0.994x, **Figure 2c**), with a mean difference of 0.21 (σ = 0.57). There is also no significant difference between the CADD scores of B/LB and VUS variants (Mann-Whitney U P=0.994. Additionally, among 125 variant pairs with SpliceAI annotations, 100.0% (N=125) did not have discordant SpliceAI scores (**Figure 3c**). This suggests the potential utility of such evidence in the variant assessment process.

Potential for aggregating VUS cases causing the same substitution: Variants may be classified as VUS based on insufficient numbers of independent cases. We sought to evaluate the frequency that two distinct sequence variants which lead to the same amino acid substitution were each classified as VUS. Clinical labs could potentially consider such additional case observations when assessing both of these variants. We note that the ACMG/AMP guidelines do not detail such associations to be evidence of pathogenicity, but we analyze the potential for aggregating cases which cause the same amino acid substitution. Overall, we find 3,790 VUS (98.5% missense / 1.5% nonsense) which are in the presence of another VUS which encodes the exact same amino acid substitution while not being in the presence of any P/LP or B/LB variants in the same codon, representing 1,985 unique VUS-VUS variant pairs affecting 1,865 distinct codons. These VUS-VUS variant pairs are often submitted by more than 2 labs in total (mean of 3.3 and a standard deviation of 2.1, **Supplementary Figure 1**), indicating that aggregating this evidence may provide substantial new evidence in favor of pathogenicity.

### Variants which lead to distinct amino acid substitutions in the same codon

When considering distinct amino acid substitutions in the same codon (PM5), we find 141,195 single nucleotide variants (excluding nonsense variants) spanning 64,373 unique codons. Among the 10,049 codons which have at least one missense variant which has been previously classified as P/LP, only 38.6% (N=3,881) are classified exclusively as P/LP. The proportion of codons which contain variants classified as VUS and P/LP is larger at 59.6% (N=5,987) and the proportion of codons which contain variants classified as P/LP and B/LB is 1.4% (N=140). The proportion of codons with missense variants of all three classifications is 0.4% (N=41, **Table 1**).

In total, this translates to 7,692 unique ClinVar VUS which have moderate evidence of pathogenicity (PM5) while not being in the presence of B/LB variants in the same codon, of which 33.5% (N=2,580, 5,666 submissions) are in clinically actionable disease genes (ACMG SF v3.0).

For these ‘PM5 P/LP-VUS variant pairs’ (P/LP and VUS missense variants which encode different amino acid substitutions in the same codon, N=9,975), the CADD scores of P/LP and VUS components are correlated (R^2^=0.169, MSE=0.624, y=0.943x, **Figure 2b**), and the distance between their CADD scores is relatively low with a mean of 0.52 (σ = 0.64). Moreover, 43.3% of the VUS have Grantham distances greater than or equal to that of their associated pathogenic variants, suggesting that some may be more deleterious.

Additionally, splicing continues to explain little discordance in variant classifications. Among 9,945 P/LP-VUS variant pairs with SpliceAI annotations, 97.2% (N=9,664) did not fall in categories where the P/LP variant was predicted to have a high impact on splicing (score >0.8) when the VUS partner was predicted to have a weak or low impact on splicing (score <0.5), or where the P/LP variant was predicted to have an intermediate impact on splicing (score (0.5, 0.8]) when the VUS partner was predicted to have a weak or low impact (**Figure 3b**).

Applicability to benign classifications: When considering evidence in favor of a benign classification (which we again note is not included in the ACMG/AMP guidelines), we evaluate the case when two variants, one classified as B/LB and the other classified as VUS, encode different amino acid substitutions at the same codon, while not being in the presence of P/LP variants in the same codon. Among 6,158 missense variant pairs, we find correlation in CADD scores (R^2^=0.376, MSE=1.212, y=0.953x, **Figure 2d**), which also have a relatively low distance between one another with a mean of 0.81 (σ = 0.75). Moreover, 43.8% of the VUS have Grantham distances less than or equal to that of their associated benign variants. Finally, among 6,083 variant pairs with SpliceAI annotations, 99.3% (N=6,042) did not have discordant SpliceAI scores (**Figure 3d**). As with PS1, this suggests that evidence analogous to PM5 for the benign classification may have utility for variant interpretation.

### LOF variants in genes where LOF is known to be pathogenic

Prior work has further defined the PVS1 criterion by identifying reasons that LOF variants may not be functionally impactful (*e*.*g*., nonsense mediated decay or exon usage).^9,11^ Additionally, metrics have been created at the gene level to help identify genes which are intolerant to loss of function variation, such as the *s*_*het*_ score and gnomAD pLI score.^4,26^ Here, we explore one additional parameter for the PVS1 criterion: the proportion of LOF variants classified as pathogenic within ClinVar genes, which reflects the certainty that loss of function variation in each gene is pathogenic. Specifically, we restrict to LOF SNVs (canonical splice-affecting and nonsense, filtered using LOFTEE to remove variants not predicted to be high-confidence loss of function) and measure the proportion which are described as pathogenic in ClinVar for each gene (**Figure 4a**).

**Figure 4:**
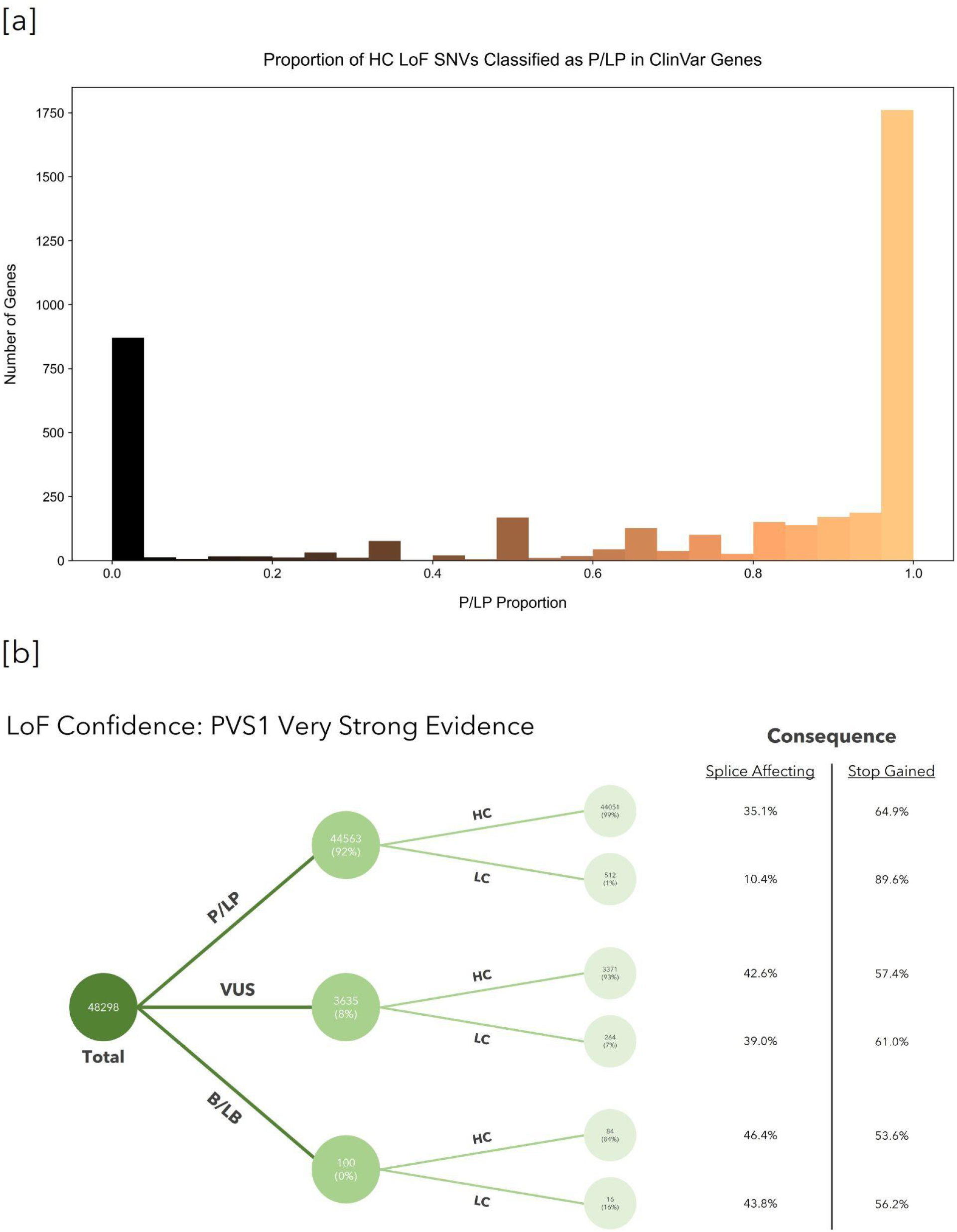
Evaluating whether LOF SNVs are predicted to be impactful in ClinVar genes. Using Loss-Of-Function Transcript Effect Estimator (LOFTEE), we classify functional impact for LOF variants (canonical splice-affecting and nonsense) as ‘Low Confidence’ (LC) or ‘High Confidence’ (HC). **[a]** For each gene in ClinVar with HC LOF variants, we measure the proportion of such variants which are pathogenic (P/LP) and find 40.1% of genes have a proportion of exactly 1.0, while 21.6% of genes have a proportion of exactly 0.0. **[b]** LOF SNV classifications in genes meeting our >50% P/LP proportion threshold. The vast majority of all LOF SNVs are estimated to be HC and impactful.

Among 4,014 genes in ClinVar which contain LOF SNVs, 2,768 have greater than 50% of the predicted high impact LOFs classified as P/LP. The 50% threshold includes a larger number of genes than would be included when filtering using high constraint (pLI) or selection (*s*_*het*_) scores (**Supplementary Figure 2**), but may be separately valuable, as ClinVar includes a broad variety of phenotypes, some of which are not under strong negative selection. When using this threshold to describe genes where LOF variants are known to be pathogenic (one possible PVS1 approach), we find 3,635 LOF variants which are classified as VUS in ClinVar, and 100 classified as B/LB, accounting for 7.5% and 0.2% of all such variants, respectively (**Table 1**). To determine whether the VUS are likely to result in loss of function, we again used LOFTEE (**Methods**) to identify any variants which might not be functionally impactful (*e*.*g*., nonsense mediated decay in last 50 bp). We find that 92.7% of LOF variants (N=3,371) are classified as ‘High Confidence’ by LOFTEE, suggesting that many of these variants are likely to be impactful (**Figure 4b**). Many LOF VUS with strong evidence of pathogenicity (PVS1) are also in clinically actionable disease genes (27.5%, N=1,000, 1,625 submissions, ACMG SF v3.0).

### Submission categories and dates of P/LP variants providing evidence of pathogenicity

Some P/LP classifications, which are based solely on assertions from literature-based sources such as OMIM, may not be based on more recent assessment criteria. However, we find that such assertions do not make up a large proportion of the associated P/LP partners of discordant VUS: only 6.6% (N=22) of P/LP variants providing PS1 evidence and 9.3% (N=931) of P/LP variants providing PM5 evidence were influenced solely by non-laboratory sources. This suggests that most discordant VUS have evidence from P/LP variants backed by laboratory submitters.

Further, as the ACMG/AMP guidelines were published in 2015, it is possible that some P/LP variants providing evidence were last evaluated on a date prior to 2015, and may not be as reliable. However, we find that only 9.8% (N=30) of P/LP variants providing PS1 evidence and 14.7% (N=1,086) of P/LP variants providing PM5 evidence were last evaluated before 2015, among variants which had an available date of last evaluation (**Supplementary Figure 3**).

It is also notable that for the PS1 and PM5 criteria, a large proportion of VUS were evaluated prior to their pathogenic partners. Among variant pairs with available dates of last evaluation, 44.7% (N=97) and 42.7% (N=2,766) of VUS were last evaluated before their pathogenic partners for the PS1 and PM5 criteria, respectively. These large percentages highlight that many discordant VUS classifications were made when evidence of pathogenicity was not known, and are likely strong candidates for reassessment, which could additionally make use of updated evidence sources (*e*.*g*., gnomAD).

### Prioritizing variants for reassessment using supporting evidence

In addition to submission dates, we provide metrics which may be useful for prioritizing variants for reassessment. These characteristics include allele frequencies (PS1, PM5, PVS1), SpliceAI scores (PS1, PM5), LOFTEE classifications (PVS1), CADD scores (PM5), and ClinVar review status [or ‘star’ rating] (PS1, PM5, PVS1).

For the PS1 criterion, we find that 43.2% (142 of 329 variant pairs with SpliceAI scores) pass all filters (**Supplementary Methods, Supplementary Table 3**). The vast majority of variants are filtered due to ClinVar review status. For PM5 evidence, this proportion drops to 28.0%, primarily due to additional filtering based on CADD scores. Finally, 25.0% of variants with PVS1 evidence of pathogenicity met all criteria.

### Identifying all nsSNVs outside of ClinVar with evidence of pathogenicity

Extending our analysis, we evaluate all nsSNVs in genes within ClinVar which could be informed by evidence of pathogenicity based on PS1, PM5, and PVS1 criteria. In total, we find over 1.4 million variants with such evidence, and provide information about their frequency and similarity to related pathogenic variants (**Supplementary Table 4, Supplementary Methods and Materials**).

### Analyzing the functional assay values of PS1 and PM5 variant pairs in BRCA1

Finally, using functional assay data from nearly 4,000 SNVs in *BRCA1*^24^, we analyze the extent to which experimental measurements of functional impact differ between missense variants in the same codon, regardless of their status in ClinVar.

First, we analyze variant pairs which encode the same amino acid substitution (PS1). We find that 98.0% of PS1 missense variant pairs have functional assay measurements that are within one functional classification (*e*.*g*., ‘functional’, ‘intermediate’, ‘loss-of-function’) of each other, 92.0% have functional scores within one standard deviation of each other, and 85.9% share the same functional classification. Overall, we observe no significant difference between functional assay value differences for PS1 variant pairs when compared with a synonymous model of differences (generated using differences in functional scores among variants causing no amino acid change, **Supplementary Figure 4**; Kolmogorov-Smirnov P=0.866). Further, we find a significant difference between PS1 variant pairs and a null model of approximately 2 million missense variant pairs which are not in the same codon (**Supplementary Figure 4**; Kolmogorov-Smirnov P=1.496×10^−26^), indicating that these score differences are significantly different from neutral expectation. This suggests that distinct variants in the same substitution often have similar measurements of functional effect, supporting the use of guidelines based on location or functional consequence. An analysis of PM5 variant pairs is included in the **Supplementary Methods and Materials**.

## Discussion

The 2015 ACMG/AMP sequence variant interpretation guidelines describe how to make use of evidence from previously classified variants during variant assessment. For the three interpretation criteria we analyzed, it appears that rules which make use of prior variant classifications may be underutilized in variant assessment. This may be because systems to review, annotate, and interpret variants often refer to nucleotide substitutions rather than amino acid substitutions. It may also relate to frequency of variant reassessment, as a substantial proportion of VUS assertions were not re-evaluated after new information became available about related variants which could have influenced variant interpretation. Other possible reasons include that there have been historically different nomenclature to describe sequence variants, or that variants within the same codon or of a related type are not identified by existing variant analysis tools.

Variant curators should be aware of how valuable these rules are and expand efforts to identify related variant data. A substantial number of SNVs are classified as uncertain, conflicting, not provided, or are outside of ClinVar, but have some form of supporting evidence from an existing classified variant in ClinVar. This information could be broadly useful for novel variant assessment by diagnostic labs, and could inform future guideline development. To make such information more accessible, we provide pre-computed lists of variants within ClinVar and exome-wide which meet these criteria (https://doi.org/10.6084/m9.figshare.c.5914178.v1).

The information could also be made more visible for individuals who search or submit to variant databases such as ClinVar. One opportunity to make this information easier to discover would be to have a section which describes ‘linked variants’, whether affecting the same codon (*e*.*g*., PS1, PM5) or having equivalent functional consequence in the appropriate context (*e*.*g*., PVS1). Separately, it may be possible to reduce variants which are submitted that have not made use of such data. While there is a process to notify a clinical submitter about a discordant classification during ClinVar submission, we note that there is no such process to notify submitters about discordant classifications or related variant data within the same codon or gene. Lines of conflicting or supporting evidence (such as the PS1, PM5, and PVS1 criteria) would be useful considerations for diagnostic labs to be made aware of during the submission process. Finally, as there are a substantial number of VUSs which are not re-evaluated once new evidence becomes available, there may be an opportunity to trigger re-evaluation of such variants as new structured evidence arises, as described in the recent AMP position statement on variant data sharing for public database operators.^27^

Standardizing how and when to apply the PVS1 criterion could be helpful to improve the utility and adherence to this form of evidence. Important work has been done to further define the PVS1 criterion by identifying LOF variants less likely to be causal and applying gene metrics which may indicate strong likelihood of variant pathogenicity. We have explored another approach based on the proportion (and certainty) of loss of function variation being classified as pathogenic in each gene. Future work may seek to identify the most appropriate threshold for this proportion, and the extent to which such data may be supported by hypothesis-free functional assays or clinical data.

Additionally, while the ACMG/AMP guidelines do not have equivalent tiers of evidence in favor of benign classification for PS1 and PM5 criteria, it appears that computational predictions of variant effects, both for VUS and variants outside of ClinVar, are similar to those of their B/LB partners to a degree that it is worth evaluating such rules in the future. If sufficiently strong, this evidence could help reduce the number of individuals who would learn about carrying a VUS in a clinically actionable disease gene, avoiding potential overtreatment and unnecessary surveillance.

Limitations: Although we have identified many occurrences of variants with uncertain, conflicting, or not provided classifications which have evidence of pathogenicity, we emphasize that these variants are not necessarily incorrectly classified, and may be influenced by other evidence. While lab submissions increasingly include text descriptions of evidence used in the classification process, it is challenging to know which criteria were specifically applied without structured data. Thus, we do not know how many discordant VUS could have an updated classification (*e*.*g*., VUS to P/LP) specifically because of the application of PS1, PM5, or PVS1 criteria. Additionally, although they comprise a small proportion, some variants providing evidence of pathogenicity are non-laboratory classifications (*e*.*g*., OMIM) and/or were last evaluated prior to 2015. Finally, the use of analogous criteria for the benign classification (mentioned previously) and the aggregation of VUS cases caused by the same substitution are not currently described in the ACMG/AMP guidelines. Future work may seek to expand on these limitations.

Overall, we believe that our tool Tocayo (a pipeline to identify related and discordant variants) and pre-computed variant lists can improve variant interpretation accuracy and efficiency.

## Data Availability

A pipeline which can be used to analyze variants from ClinVar and dbNSFP, as well as relevant input files, are provided online (https://github.com/cassalab/tocayo). Pre-computed lists of variants within ClinVar and exome-wide which meet PS1, PM5, and PVS1 criteria can be found here (https://doi.org/10.6084/m9.figshare.c.5914178.v1).

https://doi.org/10.6084/m9.figshare.c.5914178.v1

## Acknowledgements

We gratefully acknowledge support from the National Human Genome Research Institute (R01HG010372; V.B., I.A.A., J.D.F., M.L., C.A.C.) and for manuscript comments and helpful suggestions from Dr. Melissa Landrum (NCBI) and Dr. Natasha Strande (Geisinger).

## Author Contributions

Data curation: V.B.; Formal analysis: V.B.; Software: V.B., I.A.A., J.D.F.; Writing-original draft: V.B., C.A.C.; Writing-review & editing: V.B., C.A.C., M.L., I.A.A., J.D.F.

## Ethics Declaration

This study does not involve human subjects, live vertebrates, higher invertebrates, or any individual-level data. All data used were publicly available and aggregated at the variant level.

## Supplementary Methods and Materials

### Data processing and labeling

Using the ClinVar variant summary file (dated 12/19/2021), we restricted to single nucleotide variants (SNVs) in assembly GRCh38 and calculated functional consequences using VEP v105. All amino acid substitutions of moderate or high impact were considered for PS1 and PM5 criteria, and LOF variants (functional consequences of ‘stop_gained’, ‘splice_donor_variant’, ‘splice_acceptor_variant’) for the PVS1 criterion. Variants were matched by transcript for functional effects when a variant fell within more than one transcript or gene.

Variants with clinical significance values of ‘Pathogenic’, ‘Likely pathogenic’, or ‘Pathogenic/Likely pathogenic’ were labeled P/LP; ‘Benign’, ‘Likely benign’, or ‘Benign/Likely benign’ were labeled B/LB, and ‘Uncertain significance’, ‘Conflicting interpretations of pathogenicity’, and ‘not provided’ were labeled as VUS.

### Computational Score, Allele Frequency, and Other Data

All RefSeq annotations, computational predictions (CADD, PolyPhen2, SIFT), gnomAD allele frequencies (N=125,748 exomes), and SpliceAI scores (masked) were obtained from VEP v105.

### Identifying all possible related nsSNVs within ClinVar

To identify all possible variants within ClinVar which might have ACMG/AMP evidence, we evaluated every nsSNV. For PS1 and PM5 criteria, variants were mapped using matching RefSeq transcripts, and if no matching transcript ID was found due to annotation inconsistencies, any transcript with a moderate or high impact was used with a priority for canonical transcripts. ClinVar variants with no moderate or high impact transcripts in VEP, as well as those in the presence of conflicting evidence in the same codon, were filtered. For PVS1 criteria, the annotation process was the same but, instead of impact, required transcripts with a consequence of either ‘splice_donor_variant’, ‘splice_acceptor_variant’, or ‘stop_gained’.

### Identifying all possible related nsSNVs in the coding genome

To identify all possible variants which might have ACMG/AMP forms of evidence, regardless of whether they are already present in ClinVar, we enumerated all possible nsSNVs in dbNSFP v4.2a with potential PS1, PM5, or PVS1 evidence. For PS1 and PM5, this specifically included any variant which affects the same codon as an existing ClinVar variant, and for PVS1, included any essential splice site or nonsense variant. We filtered any variants which have inconsistent transcripts between dbNSFP (Ensembl) and ClinVar (RefSeq). For PS1 or PM5 evidence, all dbNSFP variants were matched to the RefSeq transcript of the ClinVar variant. Otherwise, or when no matching transcript was found, transcripts with moderate or high impact were used with a priority for canonical transcripts. Similar to ClinVar, dbNSFP variants without a transcript meeting these conditions or which were in the presence of conflicting evidence in the same codon were removed from datasets. For the PVS1 criterion, transcripts were matched based on having a matching consequence (e.g., ‘splice_donor_variant’, ‘splice_acceptor_variant’, or ‘stop_gained’), with a priority for canonical transcripts.

### Amino acid substitution and matching of LOF variants

For the PS1 criterion, variants are considered to have the same amino acid substitution and form a ‘variant pair’ if they meet either of the following criteria:

- The variants have matching amino acid substitutions in ClinVar/dbNSFP and VEP, matching RefSeq transcripts (from VEP), and are in the same gene, regardless of assembly positions.
- The variants have the same amino acid substitution in ClinVar/dbNSFP and VEP, and have matching assembly positions in the same frame, and are in the same gene, regardless of transcript.

For PM5 evidence, this criterion is modified, checking for variants which have the same amino acid position in ClinVar/dbNSFP and VEP but do not have the same amino acid substitution.

For PVS1 evidence, any stop gain (Ter/X) or canonical splice site variant (−1, -2, +1, or +2 base pairs from splice junction) is included if it is within a gene where >50% of such variants annotated as ‘High Confidence’ by LOFTEE (Loss-Of-Function Transcript Effect Estimator) are classified as P/LP in ClinVar.

Assembly positions are determined to be in the same frame based on codon position and strand direction.

### ACMG SF v3.0 analysis

We analyze VUS which are in genes specified for review of secondary findings (ACMG SF v3.0) to identify a subset in these clinically actionable disease genes. For this analysis, we filter to the 72 unique genes specified (excluding *HFE* because p.Cys282Tyr was not part of any set), and include any VUS with evidence of pathogenicity.

### Evaluation of ClinVar submission dates

Dates of last evaluation were analyzed for P/LP variants providing evidence of pathogenicity as well as their VUS partners. When checking for submissions before 2015, only the year of submission was considered, regardless of month or day, while the exact date was considered when comparing the submission dates of related P/LP and VUS variants. A substantial number of variants, especially those with PS1 evidence, had missing annotations for the date of last evaluation. Any variant pairs with missing annotations were removed.

### Annotations for prioritization of variant reassessment

When prioritizing VUS with the strongest evidence, we use various metrics which provide supporting context. These characteristics include the following:

- Allele frequencies less than 10^−3^; i.e., the P/LP variant pair component has an allele frequency less than 10^−3^ and the VUS variant pair component has an allele frequency less than 10^−3^ (PS1, PM5, PVS1).
- SpliceAI scores which are not discordant; i.e., the SpliceAI score of the P/LP variant pair component is less than or equal to 0.5 when the SpliceAI score of the VUS variant pair component is less than or equal to 0.5 (PS1, PM5).
- LOFTEE classification of ‘High Confidence’ [HC] with no flags (PVS1).
- CADD scores within one standard deviation of each other; i.e., the absolute value of the difference in CADD scores between the P/LP variant pair component and the VUS variant pair component is less than or equal to the standard deviation of the distribution of score differences when restricted to variant pairs which meet the filtering criteria based on allele frequencies and SpliceAI scores (PM5).
- Star rating of the VUS variant pair component is less than two (PS1, PM5, PVS1) and the star rating of the P/LP variant pair component is not zero (PS1, PM5).
- Star rating of the VUS variant pair component is zero (PS1, PM5, PVS1) or the star rating of the P/LP variant pair component is greater than or equal to two (PS1, PM5).

For variants with evidence analogous to PS1 and PM5 for the benign classification, analogous criteria are checked with the exception of the allele frequency criteria, which is not checked, and the SpliceAI criterion, which is modified to check for variant pairs in which the B/LB component has a score greater than 0.5 when the VUS component has a score greater than 0.5.

All filters are cumulative, meaning each subsequent filtering step is applied on the set of variants which passed all previous criteria.

### Frequency and similarity of related non-synonymous variants outside of ClinVar

We find 11,981 unique variants outside of ClinVar (42.9% missense / 57.1% nonsense) which have the same amino acid substitution as a P/LP variant in ClinVar while not being in the presence of B/LB variants in the same codon (PS1), translating to 11,186 unique sites. We also find 104,980 unique variants outside of ClinVar, excluding nonsense variants, which have a different amino acid substitution in the same codon as a missense P/LP variant in ClinVar [in the absence of B/LB variants in the codon] (PM5), translating to 28,251 unique sites. Finally, there are 1,295,274 unique nsSNV LOF variants (including nonsense and canonical splice site variants) outside of ClinVar which are in genes where more than 50% of all predicted ‘High Confidence’ LOF variants are classified as P/LP (**Supplementary Table 4**).

These variants also have similar predicted impacts: CADD scores of paired P/LP and VUS variants are highly correlated (R^2^=0.973, MSE=0.156, y=0.997x) and are not significantly different (Mann-Whitney U P=0.361) for PS1 evidence. All possible variant pairs meeting the PM5 criterion (N=109,628) show a weaker correlation, and the groups are significantly different from one another (R^2^=0.284, MSE=0.582, y=0.940x, Mann-Whitney U P=0.0). With respect to splicing impacts, 98.2% of PS1 variant pairs (N=11,756) and 97.9% of PM5 variant pairs (N=106,889) did not have discordant SpliceAI scores, among variants pairs with SpliceAI annotations. B/LB-VUS variant pairs showed similar or higher numbers for correlation (R^2^=0.923, MSE=0.150, y=0.974x), similarity (Mann-Whitney U P=0.529), and splicing (99.5%) for PS1, as well as for PM5 (R^2^=0.402, MSE=1.094, y=0.971x, Mann-Whitney U P=0.0, 99.4%). LOF variants with PVS1 evidence were overwhelmingly classified as ‘High Confidence’ by LOFTEE, at a rate of 98.0%.

### Identifying associations in BRCA1 functional data

To analyze the extent to which variants within the same codon differ from one another, we used a dataset of 3,893 variants in *BRCA1* and their associated functional scores. From this dataset, variant pairs were formed to represent PS1, PM5, synonymous, and null models.

A variant pair was included in the PS1 model if two missense variants encoded the same amino acid substitution, and a variant pair was included in the PM5 model if two missense variants encoded distinct amino acid substitutions in the same codon. Variant pairs were included in the synonymous model if two distinct variants encoded the same amino acid substitution as the reference. For the null model, all unique combinations of two missense variants were included if they were not already part of another model (PS1, PM5, or synonymous). Transcripts and assembly positions were ignored in all matching processes.

Differences in functional scores were calculated by taking the absolute value of the difference between the mean function scores of variant pair components. The functional classes of ‘functional’, ‘intermediate’, and ‘non-functional’ were defined by the study authors as functional scores greater than -0.748, between -0.748 and -1.328, and less than -1.328, respectively. The classes represent scores predicted to not disrupt function (functional), disrupt function (non-functional), and in between (intermediate).

When reviewing variant pairs which encode different amino acid substitutions in the same codon (PM5), 85.7% of all PM5 missense variants pairs had functional scores within one functional classification of each other, 78.6% had functional scores within one standard deviation of each other, and 73.1% shared the same functional classification. Compared to a null distribution of functional scores, we also find the distribution of PM5 variant functional scores to be significantly different (**Supplementary Figure 4**; Kolmogorov-Smirnov P=7.441×10^−55^).

### Statistical tests and data processing

Linear regressions and related statistics (R^2^, MSE) were produced using Scikit-learn v0.23.2. Statistical tests including the Mann-Whitney U and Kolmogorov-Smirnov tests were calculated using SciPy v1.7.3. Figures were built using Matplotlib v3.3.2, Seaborn v0.11.0, and PowerPoint 2019. Other data processing used Pandas v1.1.4, NumPy v1.21.1, Sqlite v3.30.0, Natsort v8.0.2, and Python v3.8.5.

## Supplementary Tables

**Supplementary Table 1:** Clinical significance values for discordant VUS. We include variants with clinical significance values of ‘Uncertain significance’, ‘Conflicting interpretations of pathogenicity’, or ‘not provided’ as VUS. The majority of VUS across all evidence types were of the clinical significance ‘Uncertain significance’. ^*a*^Form of evidence not described in ACMG/AMP guidelines.

| Evidence of Impact | Classification | Total VUS | Uncertain Significance | Conflicting Interpretations | Not Provided |
| --- | --- | --- | --- | --- | --- |
| PS1 | P/LP | 323 | 177<br>(54.8%) | 68<br>(21.1%) | 78<br>(24.1%) |
| PM5 | P/LP | 7,692 | 5,516<br>(71.7%) | 1,426<br>(18.5%) | 750<br>(9.8%) |
| PVS1 | P/LP | 3,635 | 2,336<br>(64.3%) | 712<br>(19.6%) | 587<br>(16.1%) |
| PS1<sup>a</sup> | B/LB | 125 | 92<br>(73.6%) | 28<br>(22.4%) | 5<br>(4.0%) |
| PM5<sup>a</sup> | B/LB | 5,960 | 5,161<br>(86.6%) | 694<br>(11.6%) | 105<br>(1.8%) |

**Supplementary Table 2:**
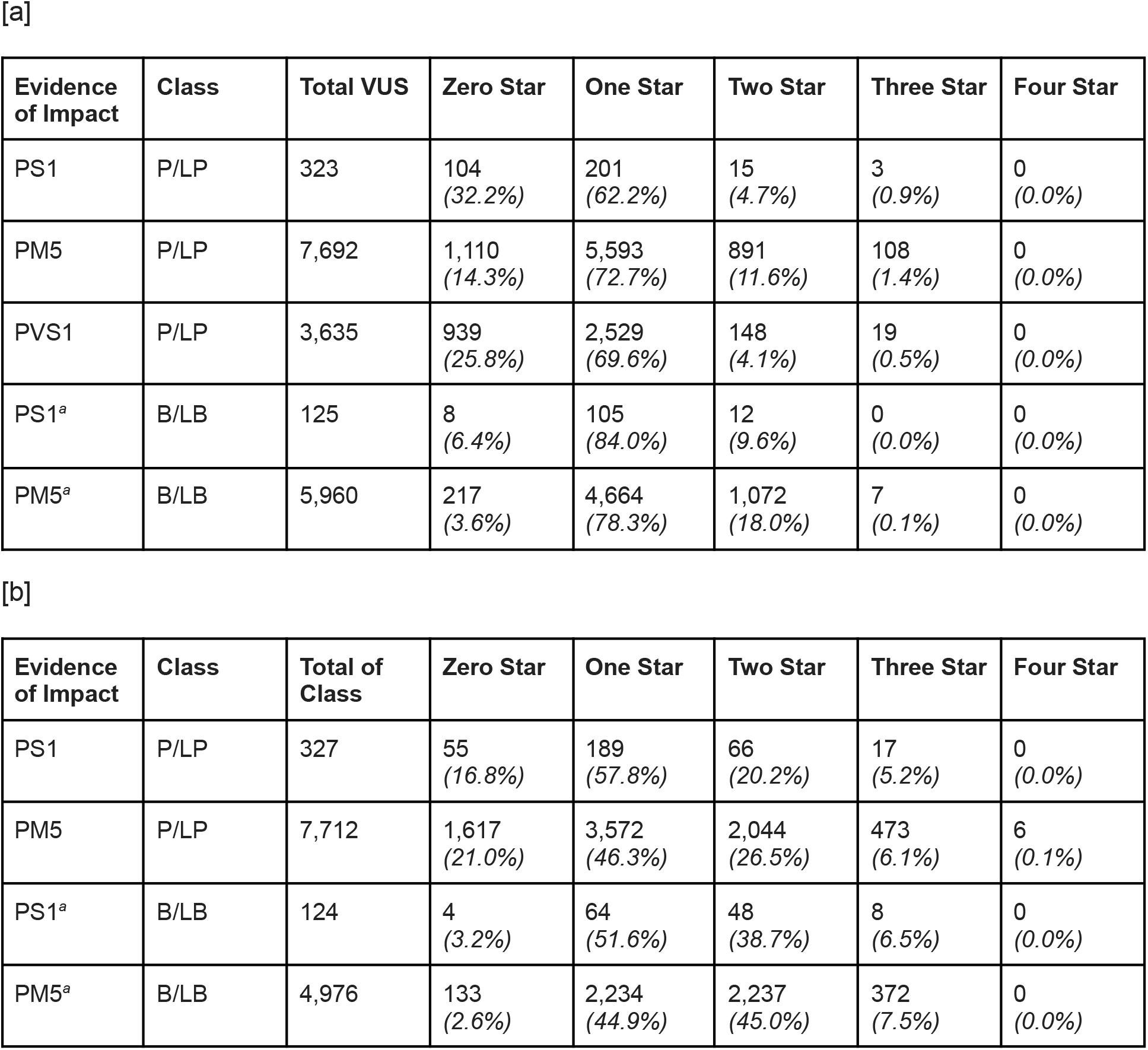
Review status of VUS, P/LP, and B/LB variants (‘star ratings’). A star rating of zero represents an incomplete interpretation, no assertion criteria and evidence provided, or not directly interpreted. A star rating of one represents a single submitter with criteria and evidence provided or multiple submitters who provided criteria and evidence but had conflicting interpretations. A star rating of two represents two or more submitters who provided criteria and evidence and had the same interpretation. A star rating of three represents an assertion reviewed by an expert panel. A star rating of four represents practice guideline. **[a]** Number of variants with each star rating for VUS with PS1, PM5, and PVS1 evidence. **[b]** Number of variants with each star rating for P/LP and B/LB variants which provide evidence to the VUS from [a]. The star ratings of the P/LP and B/LB variants are generally higher than those of their associated VUS. ^*a*^Form of evidence not described in ACMG/AMP guidelines.

| Evidence of Impact | Class | Total VUS | Zero Star | One Star | Two Star | Three Star | Four Star |
| --- | --- | --- | --- | --- | --- | --- | --- |
| PS1 | P/LP | 323 | 104<br>(32.2%) | 201<br>(62.2%) | 15<br>(4.7%) | 3<br>(0.9%) | 0<br>(0.0%) |
| PM5 | P/LP | 7,692 | 1,110<br>(14.3%) | 5,593<br>(72.7%) | 891<br>(11.6%) | 108<br>(1.4%) | 0<br>(0.0%) |
| PVS1 | P/LP | 3,635 | 939<br>(25.8%) | 2,529<br>(69.6%) | 148<br>(4.1%) | 19<br>(0.5%) | 0<br>(0.0%) |
| PS1<sup>a</sup> | B/LB | 125 | 8<br>(6.4%) | 105<br>(84.0%) | 12<br>(9.6%) | 0<br>(0.0%) | 0<br>(0.0%) |
| PM5<sup>a</sup> | B/LB | 5,960 | 217<br>(3.6%) | 4,664<br>(78.3%) | 1,072<br>(18.0%) | 7<br>(0.1%) | 0<br>(0.0%) |

**Supplementary Table 2: Review status of VUS, P/LP, and B/LB variants ('star ratings'). A**
| Evidence of Impact | Class | Total of Class | Zero Star | One Star | Two Star | Three Star | Four Star |
| --- | --- | --- | --- | --- | --- | --- | --- |
| PS1 | P/LP | 327 | 55<br>(16.8%) | 189<br>(57.8%) | 66<br>(20.2%) | 17<br>(5.2%) | 0<br>(0.0%) |
| PM5 | P/LP | 7,712 | 1,617<br>(21.0%) | 3,572<br>(46.3%) | 2,044<br>(26.5%) | 473<br>(6.1%) | 6<br>(0.1%) |
| PS1<sup>a</sup> | B/LB | 124 | 4<br>(3.2%) | 64<br>(51.6%) | 48<br>(38.7%) | 8<br>(6.5%) | 0<br>(0.0%) |
| PM5<sup>a</sup> | B/LB | 4,976 | 133<br>(2.6%) | 2,234<br>(44.9%) | 2,237<br>(45.0%) | 372<br>(7.5%) | 0<br>(0.0%) |

**Supplementary Table 3:** Number of variants by evidence with cumulative filters. All numbers are among variant pairs with SpliceAI annotations. **[a]** P/LP-VUS variant pairs with PS1 and PM5 evidence and variants with PVS1 evidence cumulatively filtered by relevant criteria. Filters regarding the star rating of the P/LP variant do not apply to the PVS1 column. **[b]** B/LB-VUS variant pairs with PS1 and PM5 evidence cumulatively filtered by relevant criteria. ^*a*^Form of evidence not described in ACMG/AMP guidelines.

| Criteria (P/LP) | PS1 | PM5 | PVS1 |
| --- | --- | --- | --- |
| None | 329 | 9,945 | 3,635 |
| Allele frequencies $<10^{-3}$ | 329 | 9,903 | 3,614 |
| & SpliceAI score of P/LP variant not $>0.5$ when VUS $\leq 0.5$ | 325 | 9,622 | - |
| & LOFTEE classification of HC as well as no flags | - | - | 3,270 |
| & CADD scores within 1 std. deviation of each other | - | 7,174 | - |
| & Star rating of VUS $<2$ and P/LP variant star rating not 0 | 261 | 5,206 | 3,124 |
| & Star rating of P/LP variant $\geq 2$ or star rating of VUS is 0 | 142 | 2,786 | 908 |

Supplementary Table 3: Number of variants by evidence with cumulative filters. All
| Criteria (B/LB)<sup>a</sup> | PS1 | PM5 |
| --- | --- | --- |
| None | 125 | 6,083 |
| & SpliceAI score of B/LB variant not $\leq 0.5$ when VUS $>0.5$ | 125 | 6,042 |
| & CADD scores within 1 std. deviation of each other | - | 3,534 |
| & Star rating of VUS $<2$ and B/LB variant star rating not 0 | 110 | 2,833 |
| & Star rating of B/LB variant $\geq 2$ or star rating of VUS is 0 | 52 | 1,526 |

**Supplementary Table 4:** Number of unique sites outside of ClinVar by evidence and classification. For PS1 and PM5, sites refer to unique codons, while for PVS1, they refer to unique variants. PS1 and PM5 sites exclude variants outside of ClinVar which are in the presence of both P/LP and B/LB variants in the same codon. ^*a*^Form of evidence not described in ACMG/AMP guidelines.

| Classification | PS1 Sites | PM5 Sites | PVS1 Sites |
| --- | --- | --- | --- |
| P/LP | 11,186 | 28,251 | 1,295,274 |
| B/LB<sup>a</sup> | 5,247 | 32,851 | - |

## Supplementary Figures

**Supplementary Figure 1:**
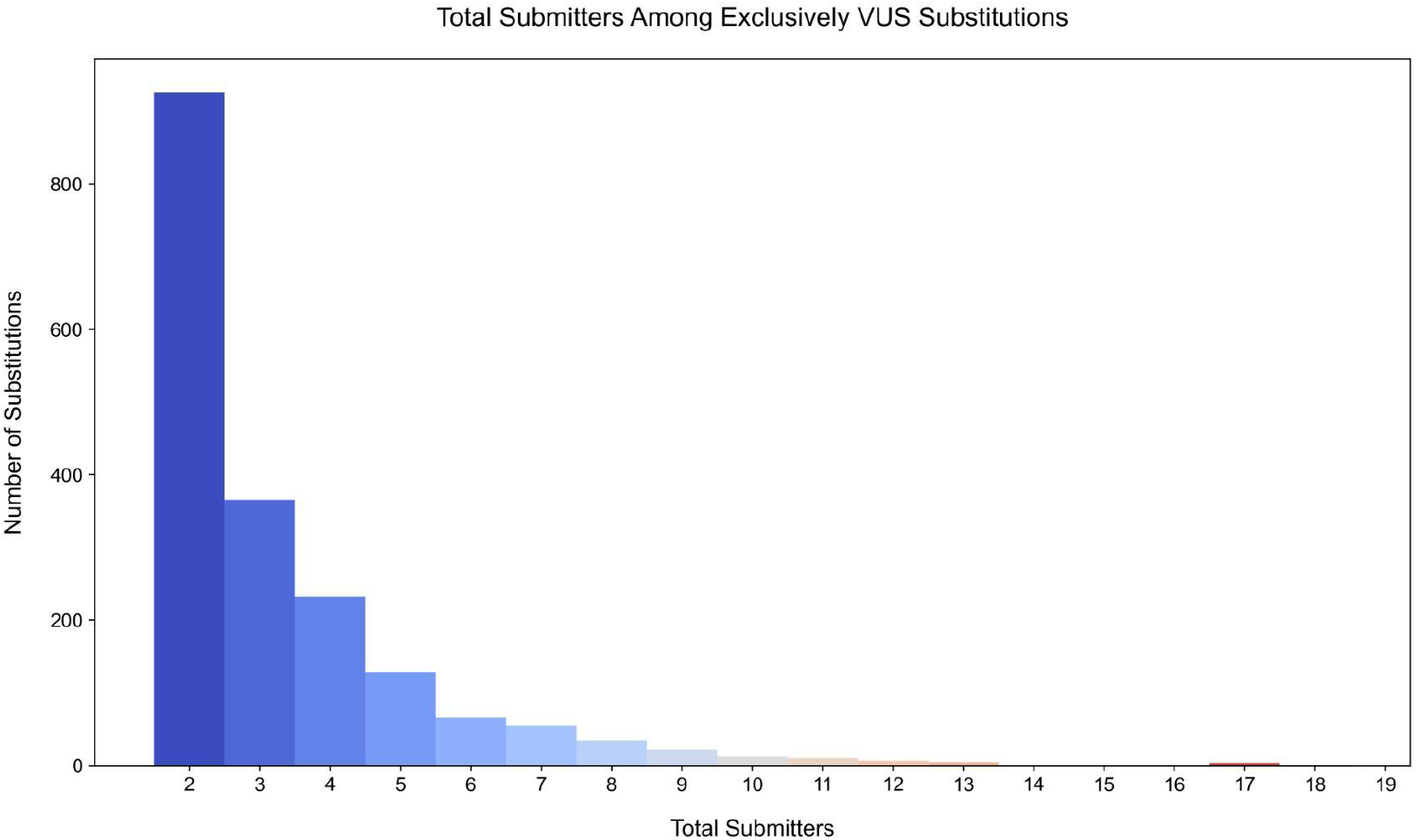
Total number of ClinVar submitters for each variant, among exclusively VUS substitutions. The sum of the number of submitters was taken for all amino acid substitutions encoded by multiple VUS while not being in the presence of any other P/LP or B/LB variants in the same codon. The distribution of these sums is strongly right skewed. 49.7% of substitutions (N=926) have 2 submitters collectively, while 18.3% (N=342) have 5 or more submitters collectively.

**Supplementary Figure 2:**
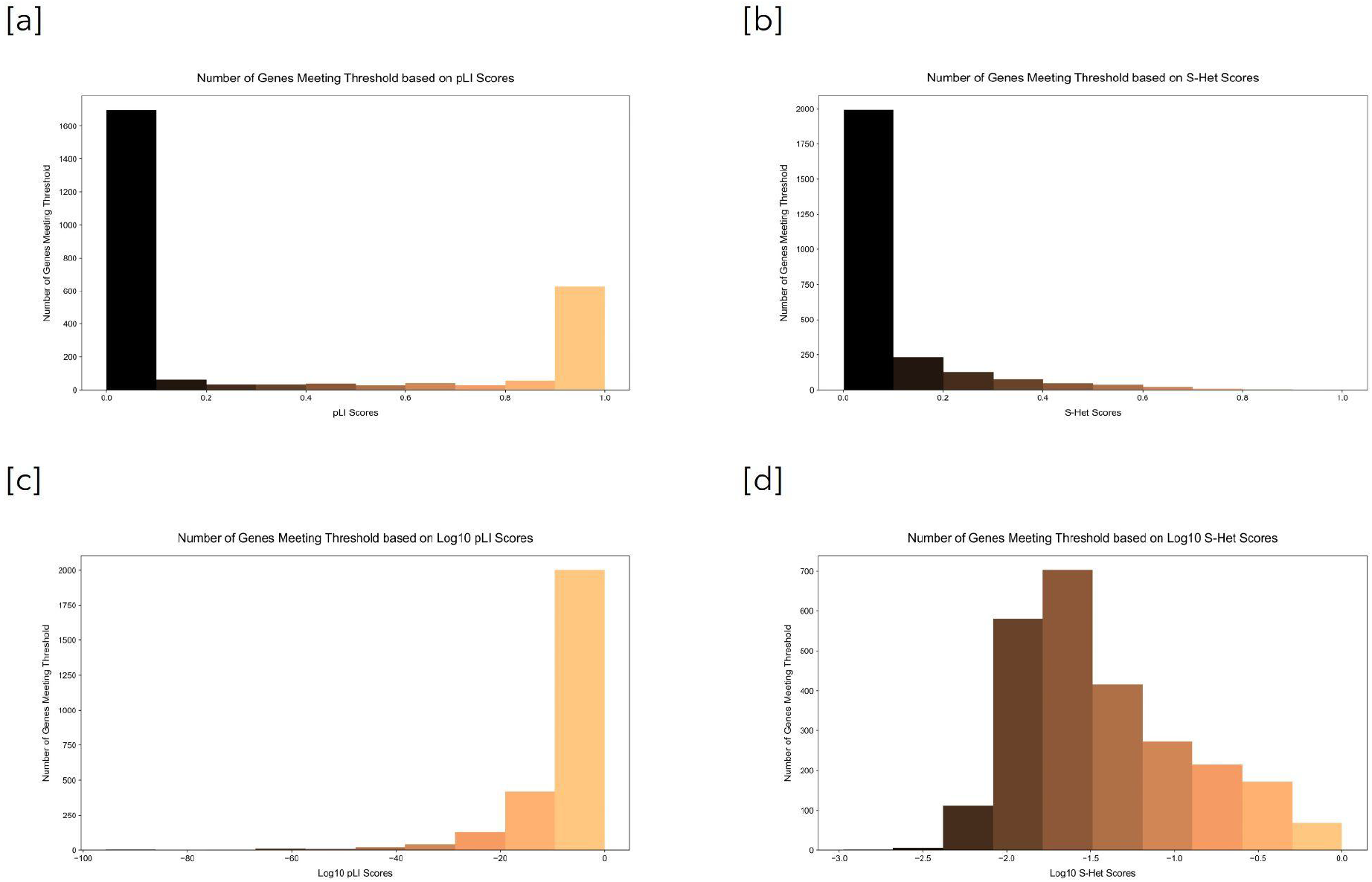
Number of genes meeting threshold based on pLI and *s*_*het*_ scores. Higher pLI and *s*_*het*_ scores represent greater intolerance to LOF variants. Genes without pLI or *s*_*het*_ scores were excluded from their respective figure panels. **[a]** Histogram of the number of genes which meet the threshold (more than 50% of ‘High Confidence’ LOF variants being P/LP in ClinVar) by pLI score deciles. **[b]** Histogram of the number of genes which meet the threshold (more than 50% of ‘High Confidence’ LOF variants being P/LP in ClinVar) by *s*_*het*_ score deciles. **[c]** Histogram of the number of genes which meet the threshold (more than 50% of ‘High Confidence’ LOF variants being P/LP in ClinVar) by log10 pLI scores. **[d]** Histogram of the number of genes which meet the threshold (more than 50% of ‘High Confidence’ LOF variants being P/LP in ClinVar) by log10 *s*_*het*_ scores.

**Supplementary Figure 3:**
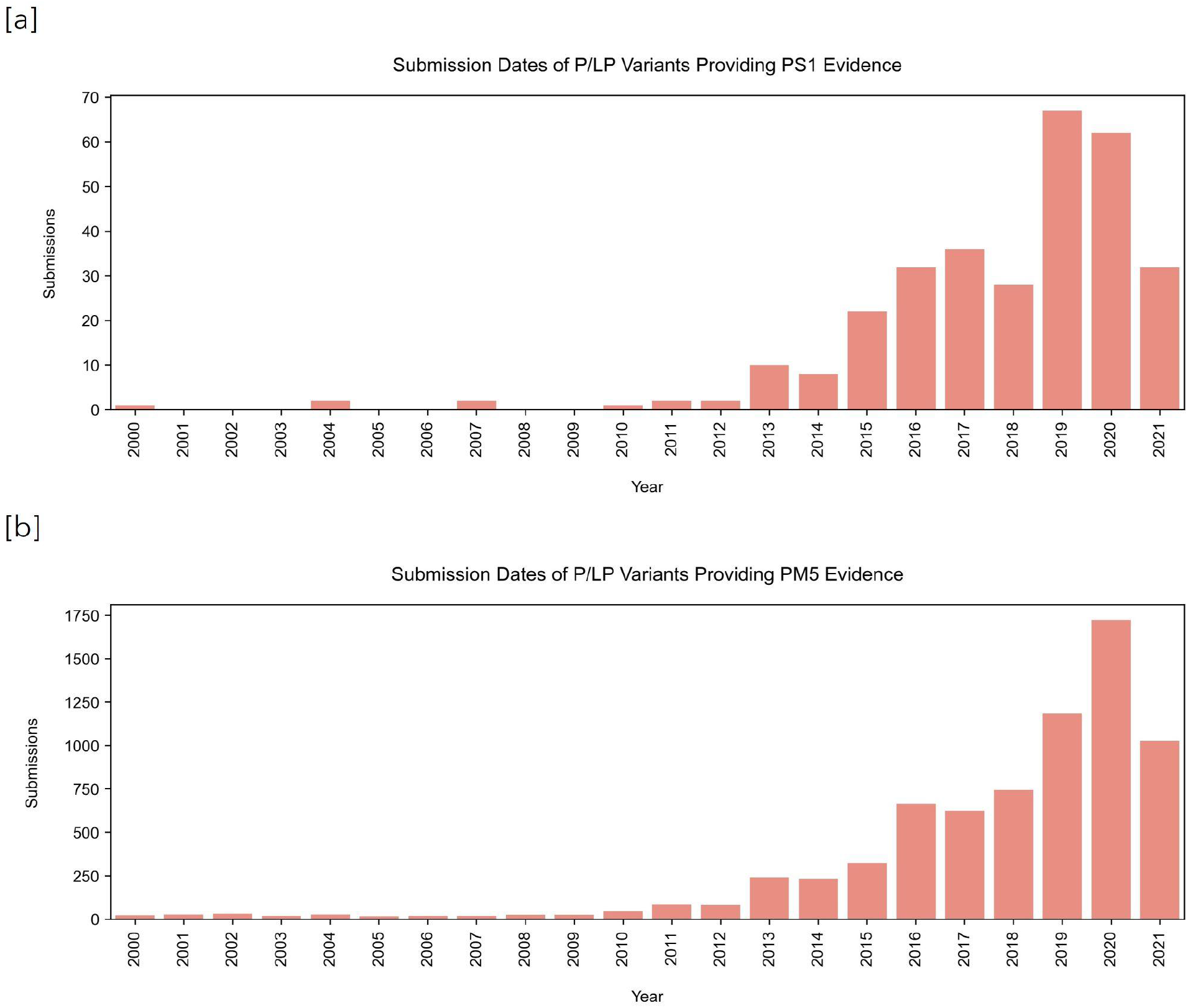
Submission dates of P/LP variants providing evidence of pathogenicity. **[a]** Submission dates of P/LP variants providing PS1 evidence. 9.8% had submission dates before 2015 when the ACMG/AMP clinical guidelines were published. **[b]** Submission dates of P/LP variants providing PM5 evidence. 14.7% had submission dates before 2015 when the ACMG/AMP clinical guidelines were published.

**Supplementary Figure 4:**
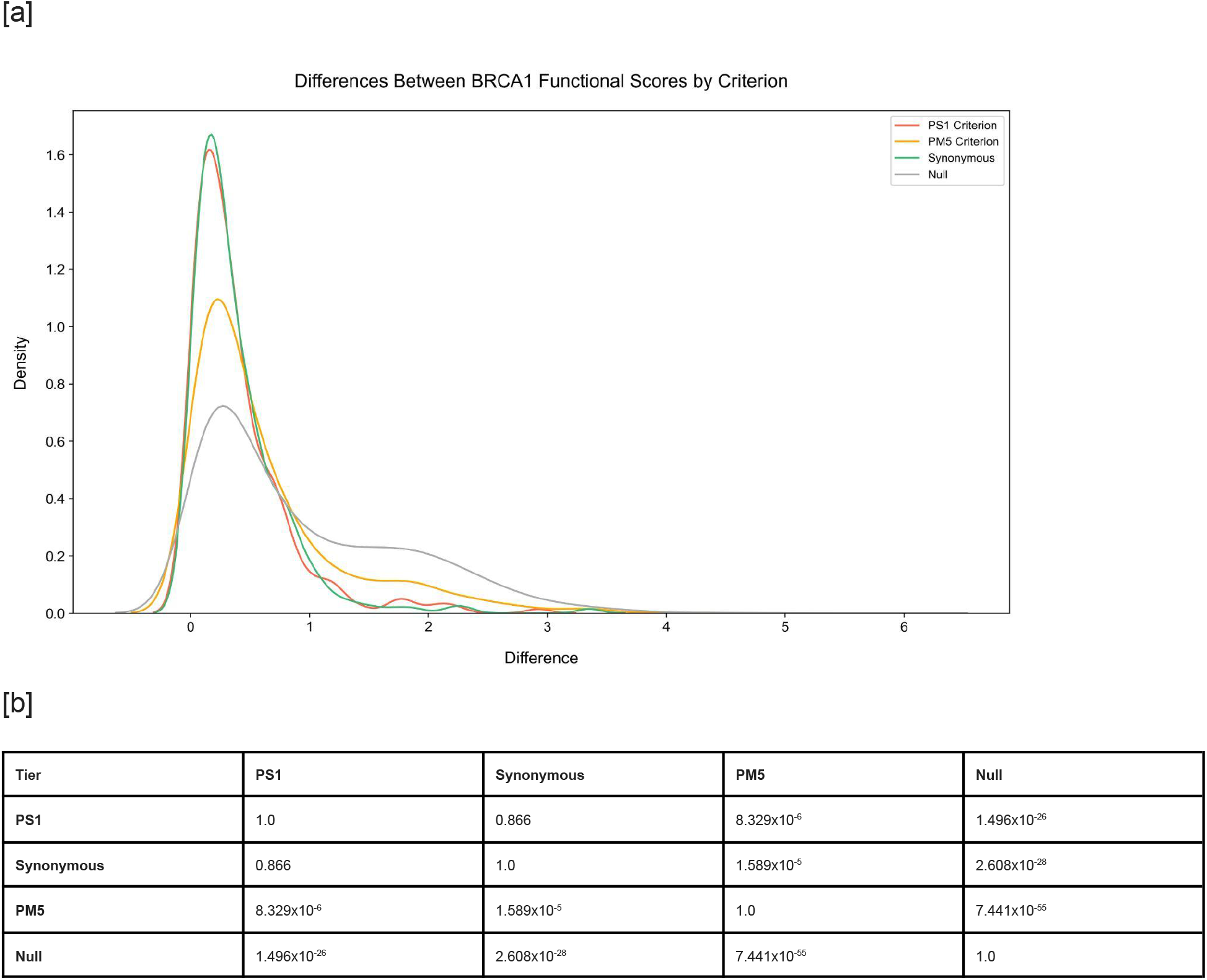
Distributions of *BRCA1* functional score differences. Of the 4 evidence tiers, PS1 (N=268) represents two missense variants encoding the same amino acid change, Synonymous (N=284) represents two synonymous variants encoding the same amino acid change, PM5 (N=1,703) represents two missense variants encoding different amino acid changes at the same position, and null (N=2,172,703) represents all possible missense variant combinations of two excluding ones included in previous tiers. All data is based on a functional study of *BRCA1* variants (N=3,893). **[a]** Density curves of the distributions of functional score differences in different evidence tiers. All distributions are right-skewed, but to varying degrees. **[b]** Kolmogorov-Smirnov p-value matrix table comparing the functional score difference distributions of different evidence tiers. The distributions of PS1 and Synonymous are most similar, while the distributions of PM5 and null are most different.

